# Two-sample Mendelian randomization analysis of circulating inflammatory markers, mammographic density phenotypes, and breast cancer subtypes

**DOI:** 10.64898/2026.09.08.26362541

**Authors:** Austin Hammermeister Suger, Tabitha A. Harrison, Cameron B. Haas, Hongjie Chen, Shaoqi Fan, Manjeet K. Bolla, Qin Wang, Joe Dennis, Kyriaki Michailidou, Alison M. Dunning, Douglas F. Easton, Melissa C. Southey, Peter A. Fasching, Lothar Haeberle, Jennifer Stone, Manuela Gago-Dominguez, Jose E. Castelao, Jennifer Ritonja, Rachel A. Murphy, D. Gareth Evans, Sherene Loi, kConFab Investigators, Fergus J. Couch, Siddhartha Yadav, Roger L. Milne, Christopher A. Haiman, Christopher G. Scott, Aaron Norman, Esther M. John, Anna Marie Mulligan, Vessela N. Kristensen, A. Heather Eliassen, Irene L. Andrulis, Jonine D. Figueroa, Susan Astley, Kamila Czene, Paul D.P. Pharoah, Antonis C. Antoniou, Montserrat García-Closas, Amy Berrington, Gretchen L. Gierach, Celine M. Vachon, Rulla M. Tamimi, Sara Lindström

## Abstract

**Background:** Observational studies have identified associations between circulating inflammatory markers (CIMs), mammographic density (MD) phenotypes, and breast cancer, but whether these associations reflect causal effects remains unclear.

**Methods:** We utilized two-sample Mendelian randomization (MR) analyses to assess relationships between 60 CIMs and three MD phenotypes, overall breast cancer, and five subtypes in individuals of European genetic ancestry by examining associations between genetically predicted CIM concentrations, three MD phenotypes (dense area, DA; non-dense area, NDA; percent density, PMD), overall breast cancer and five subtypes (ER-, HER2+, luminal A, LumA, luminal B, LumB and triple-negative, TN). Primary analyses used the inverse variance weighted (IVW) MR method.

**Results:** In IVW MR analyses, we observed associations between 4 CIMs and breast cancer subtypes (outcome/direction of association): CASP-8 (overall/+ & TN/+), CXCL9 (overall/− & LumA/−), LIF-R (overall/− & LumA/−), and MMP-10 (LumB/+) that remained significant after Bonferroni correction for multiple testing. At a nominal significance level of *p* < 0.05, we observed suggestive evidence for eight MD-CIM associations: CCL23 (DA/+), CD40 (NDA/−), GDNF (DA/+ & PMD/+), MMP-10 (DA/+ & PMD/+), TRANCE (DA/−), and TWEAK (NDA/+). In multivariable MR analyses for MD phenotypes accounting for body mass index (BMI), we observed six nominal associations represented by six unique CIMs, although most instruments demonstrated weak conditional instrument strength after inclusion of BMI.

**Conclusion:** Overall, our comprehensive analysis of 60 CIMs replicated some known breast cancer associations but provides limited evidence supporting broad causal effects of circulating inflammatory markers contributing to variation in MD phenotypes.

## Introduction

Female breast cancer is the most diagnosed cancer among women worldwide, with an estimated 2.3 million incident cases diagnosed in 2022, and remains a major contributor to cancer mortality (1). Mammographic density (MD), defined by the relative amounts of connective, epithelial, and adipose tissue in the breast, is a well-established risk factor for breast cancer (2–4). MD phenotypes include mammographic dense area (DA), non-dense area (NDA), and percentage of dense area in the breast (percent mammographic density; PMD). In addition to being a breast cancer risk factor, increased MD (higher PMD or DA) is associated with reduced sensitivity and specificity for mammography screening (5,6). Characterizing factors associated with both breast cancer and MD could help us better understand its influence on breast cancer risk and impact on screening.

Tumor-promoting inflammation is a cancer hallmark (7,8) and circulating inflammatory markers (CIMs) have been associated with breast cancer in molecular and observational epidemiological studies (9–11). One of the most studied CIMs is C-reactive protein (CRP), which has a functional role in inflammatory processes (12), with studies supporting its involvement in apoptosis, immune cell recruitment, and lipid accumulation (13). Multiple studies have observed an association between higher concentrations of circulating CRP and risk of breast cancer (14–17), yet many of the identified associations were modest and varied by body mass index (BMI), menopausal status, or other factors. Relationships between other CIMs and breast cancer risk have been studied less extensively and the associations that have been detected are often inconsistent across studies and vary by breast cancer subtype, BMI, menopausal status, menopausal hormone therapy use, or other factors (18–20).

Elucidating the relationships between CIMs, breast cancer risk factors, and breast cancer could help disentangle the etiological role of systemic inflammation in breast cancer risk. Studies have identified associations between MD phenotypes and circulating concentrations of interleukin 6 (IL-6) (21–23), interleukin 8 (IL-8) (22,23), tumor necrosis factor receptor 2 (TNFR2) (24), vascular endothelial growth factor (VEGF) (23), and CRP (21,22,24,25). However, similar to the breast cancer studies, many associations were inconsistent across studies and varied by potential confounding factors (21,25).

The objective of this study was to comprehensively assess associations between genetically predicted CIMs, MD phenotypes, and breast cancer using a two-sample Mendelian randomization (MR) approach, as MR can help mitigate some methodological limitations of observational studies, such as reverse causation and uncontrolled confounding. We further performed multivariable MR analyses for MD outcomes including genetically-predicted BMI as an exposure, given its strong association with MD phenotypes and involvement in inflammatory processes (4,26). Previous bidirectional MR studies of CRP and BMI have suggested that BMI is likely to affect CRP, rather than vice versa, and thus BMI is a potential confounder in CRP-MD phenotype relationships (27,28). In total, we conducted MR analyses of 60 genetically predicted CIM exposures in relation to three MD phenotypes, and six breast cancer outcomes.

## Methods

### MR study design and data sources

The STROBE-MR checklist was used to guide the reporting of our MR analyses (29). Causal diagrams illustrating the relationships that we investigated using MR and multivariable MR analyses are shown in Figure 1. We used summary statistics from published European genetic ancestry GWAS for BMI (*N subjects* = 461,460) (30), circulating CRP (*N* = 575,531) (31) and TNFR2 (*N* = 21,758) (32), and circulating levels of 58 additional inflammation-related proteins (*N* = 14,824) for which at least one genome-wide significant (*p* < 5×10^−8^) single nucleotide polymorphism (SNP) exists (33) (Table S1), for a total of 60 unique CIMs. GWAS summary statistics were obtained from either the IEU OpenGWAS project (34,35), the EBI-NHGRI GWAS Catalog (36), or study-specific repositories (33), selecting the largest available European genetic ancestry GWAS when multiple were available. For MD phenotypes (DA: *N* = 13,965; NDA: *N* = 14,036; PMD: *N* = 17,841), we used GWAS summary statistics from the Breast Cancer Association Consortium (BCAC) dataset described in Haas et al., 2024 (37) (Table S2). For all breast cancer outcomes except ER-(overall: *N* = 247,173; triple-negative, TN: *N* = 106,377; luminal A, LumA *N* = 148,877; luminal B, LumB *N* = 105,277; HER2+ *N* = 97,877), we used publicly available summary statistics described in Zhang et al., 2020 (38). For ER-breast cancer (*N* = 72,261), we used publically available summary statistics described in Milne et al., 2017 (39). The original GWAS or studies meta-analyzed by those GWAS obtained ethical approval from their organization/institution(s) ethical review boards as described in the cited articles. The GWAS summary statistics used did not contain any individual-level information.

**Figure 1.**
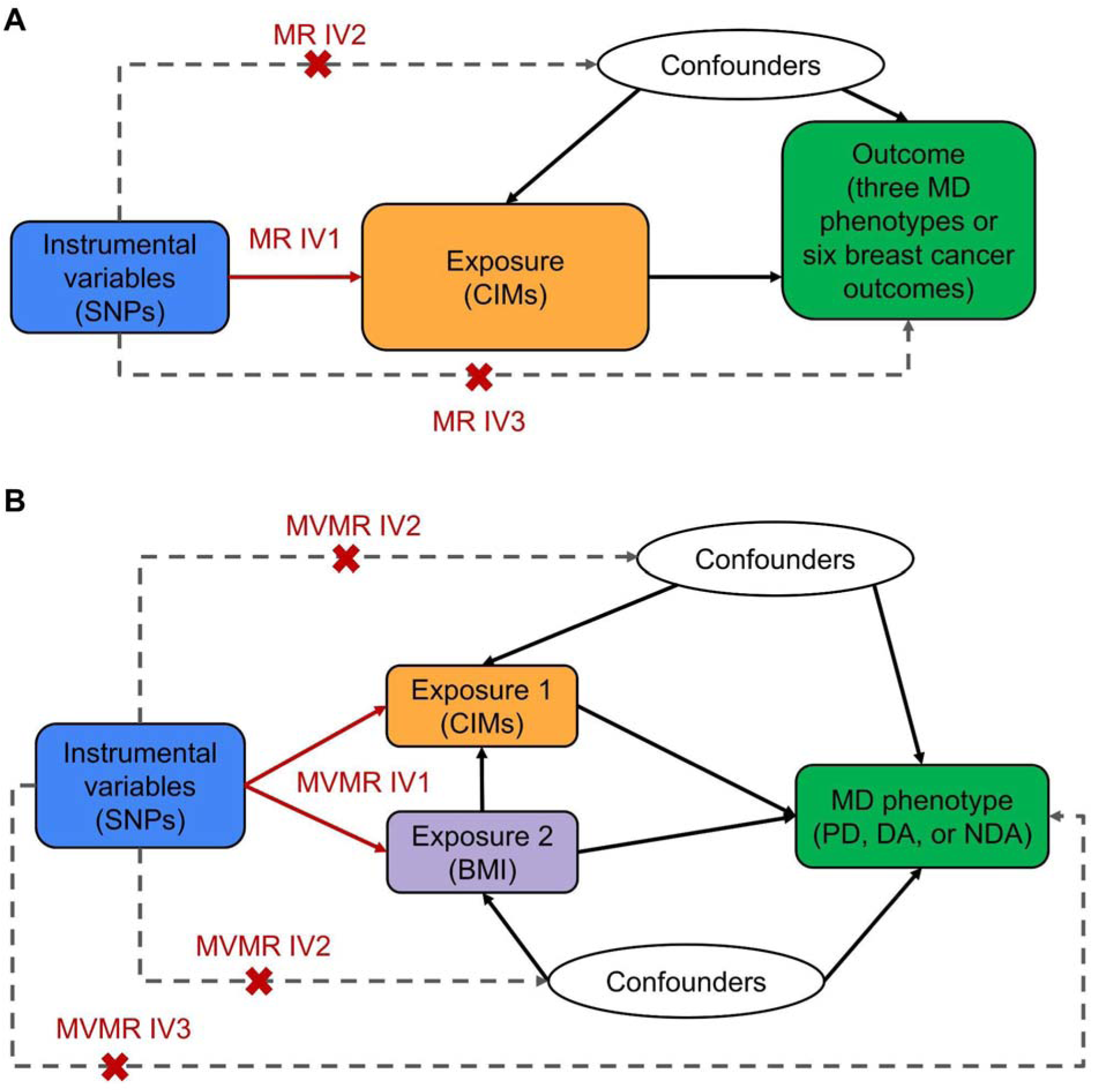
Causal diagrams representing the relationships investigated in Mendelian randomization (MR) and multivariable MR (MVMR) for mammographic density (MD) phenotypes and breast cancer outcomes with core assumptions indicated. (A) Causal diagram representing univariable MR analyses for CIM exposures and breast cancer and MD phenotype outcomes. Core MR instrumental variable (IV) assumptions (MR IV1-IV3) are highlighted in red. (B) Causal diagram representing CRP MVMR analyses for MD outcomes. Core MVMR IV assumptions (MVMR IV1-IV3) are highlighted in red.

### MR assumptions

To be valid, instrumental variables (IVs) used in MR analyses must meet three assumptions (MR IV 1-3): (1) the IVs must be strongly associated with the exposure, (2) the IVs must be independent of all (observed or unobserved) confounders of the exposure-outcome relationship, and (3) the IVs must be associated with the outcome only via the exposure (i.e., independent of the outcome given the exposure; Figure 1A) (40). The expanded assumptions (multivariable MR IV 1-3) for valid IVs in multivariable MR analyses are that: (1) the IVs must be strongly associated with each exposure given all other exposures in the model, (2) the IVs must be independent of all confounders of any of the relationships between the exposures and the outcome, and (3) the IVs must be independent of the outcome given all exposures (Figure 1B) (40,41).

### Statistical analyses

We used the TwoSampleMR R package v0.6.11 (RRID:SCR_019010) (34), custom functions in R v4.3.2 (RRID:SCR_001905) (42), and PLINK 2.0 alpha v6.5 (43,44) to extract and process exposure GWAS summary statistics, and harmonize the exposure and outcome GWAS summary statistics to create MR instruments. For each exposure GWAS, only SNPs with *p* < 5×10^−8^ were extracted, then SNPs with linkage disequilibrium (LD) were excluded (r^2^ = 0.001 and clumping window = 10,000 base-pairs) using the 1000 Genomes Project European genetic ancestry (1000G EUR) phase 3 data as an LD reference panel (34,45). For the multivariable MR instruments other than CRP, if a SNP was not included in both the CIM and BMI GWAS summary statistics, then a proxy SNP with r^2^ > 0.8 within 500 kb in the 1000G EUR LD reference panel was used (35). The closest LD proxy SNP with the highest r^2^ was always selected. Ambiguous strand information was resolved using allele frequency information when possible. Palindromic SNPs were retained if strand information could be inferred.

When constructing all instruments, we excluded SNPs with a minor allele frequency (MAF) below 5% in the 1000G EUR reference panel (45). We constructed two instruments for the CRP univariable MR analyses to try to disentangle the influences of CRP from BMI on MD phenotypes. The first instrument (CRP) included SNPs that met the standard inclusion criteria described above. The second instrument (CRP_noBMI) was further restricted to CRP-associated SNPs that did not show an association (*p* ≥ 0.05) with BMI and were not in LD with a BMI-associated SNP (r^2^ > 0.1 within 500 kb) in the 1000G EUR LD reference panel (36,45,46). The number of SNPs in each of the final instruments are shown in Table S3.

For all CIM instruments, we conducted univariable MR analyses for all outcomes and multivariable MR analyses for MD phenotype outcomes using random-effect inverse-variance weighted (IVW) methods (47). These methods were implemented in the MendelianRandomization R package v0.10.0 (RRID:SCR_025049) (48). Primary inference was based on random-effects IVW MR analyses, with nominal *p*-values providing suggestive evidence in the context of multiple testing across CIM-MD and CIM-breast cancer outcome pairs. A total of 61 CIM instruments with at least one valid SNP were tested using the univariable MR method for each of the three MD phenotypes and six breast cancer outcomes. CRP had two instruments for univariable MR analyses as described above. This resulted in 183 (61 x 3) and 366 (61 x 6) total univariable MR analyses for the MD phenotype and breast cancer outcomes, respectively. For multivariable MR, a total of 60 instruments resulted in 180 total multivariable MR analyses for 180 unique CIM-MD associations. As a result, the corresponding Bonferroni-corrected significance thresholds were *p* < 2.73 × 10^−4^ (0.05 / 183), *p* < 1.37 × 10^−4^ (0.05 / 366) and *p* < 2.78 × 10^−4^ (0.05 / 180), respectively, for univariable MD phenotypes, univariable breast cancer, and multivariable IVW MR analyses.

### Assessment of MR assumptions and sensitivity analyses

In univariable MR analyses, we calculated *F*-statistics for each of the instruments to assess the MR IV1 assumption (48). Although there are no formal tests for the MR IV2 and IV3 assumptions, we used up to seven methods with varying degrees of robustness to help identify potential violations. For instruments with at least one SNP, we calculated MR Wald ratios for each SNP in the instrument. For instruments with more than two SNPs, we further used the MR-Egger (49), MR weighted median (MR median), MR-LASSO (50), and MR constrained maximum-likelihood (MR cML) (51) methods implemented in the MendelianRandomization R package (48). For instruments with more than 10 SNPs, we used the MR pleiotropy residual sum and outlier (MR-PRESSO) method (52). The MR-LASSO, MR cML, and MR-PRESSO methods support the identification of outliers and potentially invalid SNPs in the instruments.

To assess the multivariable MR IV1 assumption, we calculated Sanderson Windmeijer conditional *F*-statistics for each exposure in the multivariable MR analyses. For the CRP multivariable MR analyses, we also incorporated the estimated phenotypic correlation between CRP and BMI in UK Biobank (UKB) participants into the conditional *F*-statistic calculations. This phenotypic correlation was estimated by Pearson correlation between CRP ln(mg/l) and BMI (kg/m^2^) at baseline for 466,356 participants with available data. To identify violations of multivariable MR IV2-IV3 and outliers or invalid SNPs, we used multivariable extensions of the MR-Egger, MR weighted median, MR-LASSO, MR-cML, and MR generalized method of moments (MR GMM) methods implemented in the MendelianRandomization R package (34). For MR cML and multivariable MR cML, the range of potentially invalid IVs was based on the number of potentially invalid IVs detected by MR-LASSO or multivariable MR-LASSO, respectively. Given the strong correlation between BMI and CRP, we used two additional multivariable MR methods for the CRP and BMI (CRP_BMI) analyses: the multivariable MR-PRESSO method implemented in the MR-PRESSO R package v1.0 (RRID:SCR_023697) (52) and robust Q-statistic minimization method implemented in the multivariable MR (MVMR) R package v0.4.1 (41).

## Results

### Study population

The SNP-exposure effects estimated in the CIM GWAS populations are expected to be similar to those in the BCAC MD phenotype and breast cancer GWAS populations, as all individuals were of inferred European genetic ancestry. None of the BCAC studies included in the MD phenotype or breast cancer GWASs were included in any of the CIM GWASs, and any sample overlap is deemed minimal (Tables S1 & S2). The estimated Pearson correlation between CRP ln(mg/l) and BMI (kg/m^2^) at baseline in UKB participants (*N* = 466,356) was r = 0.43.

### Univariable MR analysis

All instruments used in univariable MR analyses had *F*-statistics > 10, suggesting strong instruments (Table S3). All the SNPs included in the CIM instruments were either associated with that CIM or were in high LD (r^2^ > 0.8) with a SNP associated with that CIM in the GWAS Catalog or a previous publication (36,46).

In univariable random-effects IVW MR analyses, we observed associations for CASP-8 (overall and triple-negative breast cancer), CXCL9 (overall and luminal A breast cancer), LIF-R (overall and luminal A breast cancer), and MMP-10 (ER-negative breast cancer) that were significant after Bonferroni correction for multiple testing (Table 1 & Figure 2). These associations were generally directionally consistent across breast cancer subtypes (Table 1, Figure 2, & Table S4). For MD outcomes, we observed nominally significant (*p* < 0.05) associations for CCL23, GDNF, MMP-10, and TRANCE with DA, CD40 and TWEAK with NDA, and GDNF and MMP-10 with PMD (Figure 3, & Table S4). For DA, the effect estimates for CCL23 and TRANCE were directionally consistent across multiple MR sensitivity analyses (Figure 3, Tables S4-S9). Similarly, the effect estimate for TWEAK with NDA was generally consistent across sensitivity analyses (Figure 3, Tables S4 & S5). The effect estimates for GDNF and MMP-10 were directionally consistent across DA and PMD analyses (Table S4). The instruments for CASP-8, CD40, CXCL9, GDNF, LIF-R and MMP-10, each included a single SNP, precluding additional multi-SNP sensitivity analyses.

**Figure 2.**
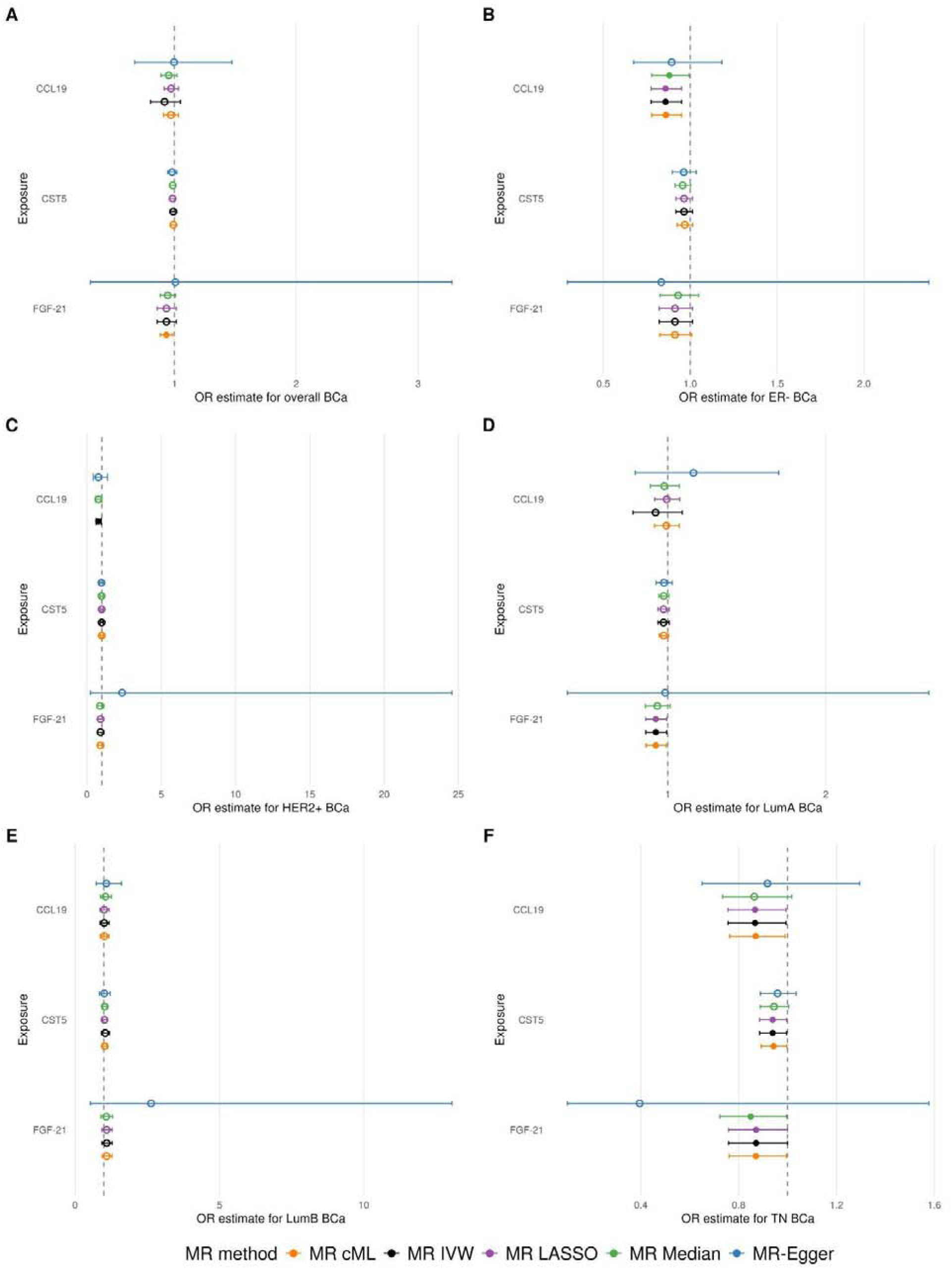
Inverse variance weighted Mendelian randomization (MR) analysis results for significant circulating inflammatory marker (CIM) instruments with at least three SNPs in breast cancer analyses. CIM instruments containing at least three SNPs and with at least one nominally significant (*p* < 0.05) association with a breast cancer outcome are listed on the y-axes. MR OR estimates and 95% confidence intervals for A) overall, B) ER-negative, C) HER2-positive, D) Luminal A, E) Luminal B, and F) triple-negative breast cancer are on the x-axes. MR methods are indicated by the point and error bar color, with the Egger, median, LASSO, IVW, and cML MR methods plotted from top to bottom on each plot for each CIM. Open circles represent MR estimates with *p* ≥ 0.05.

**Figure 3.**
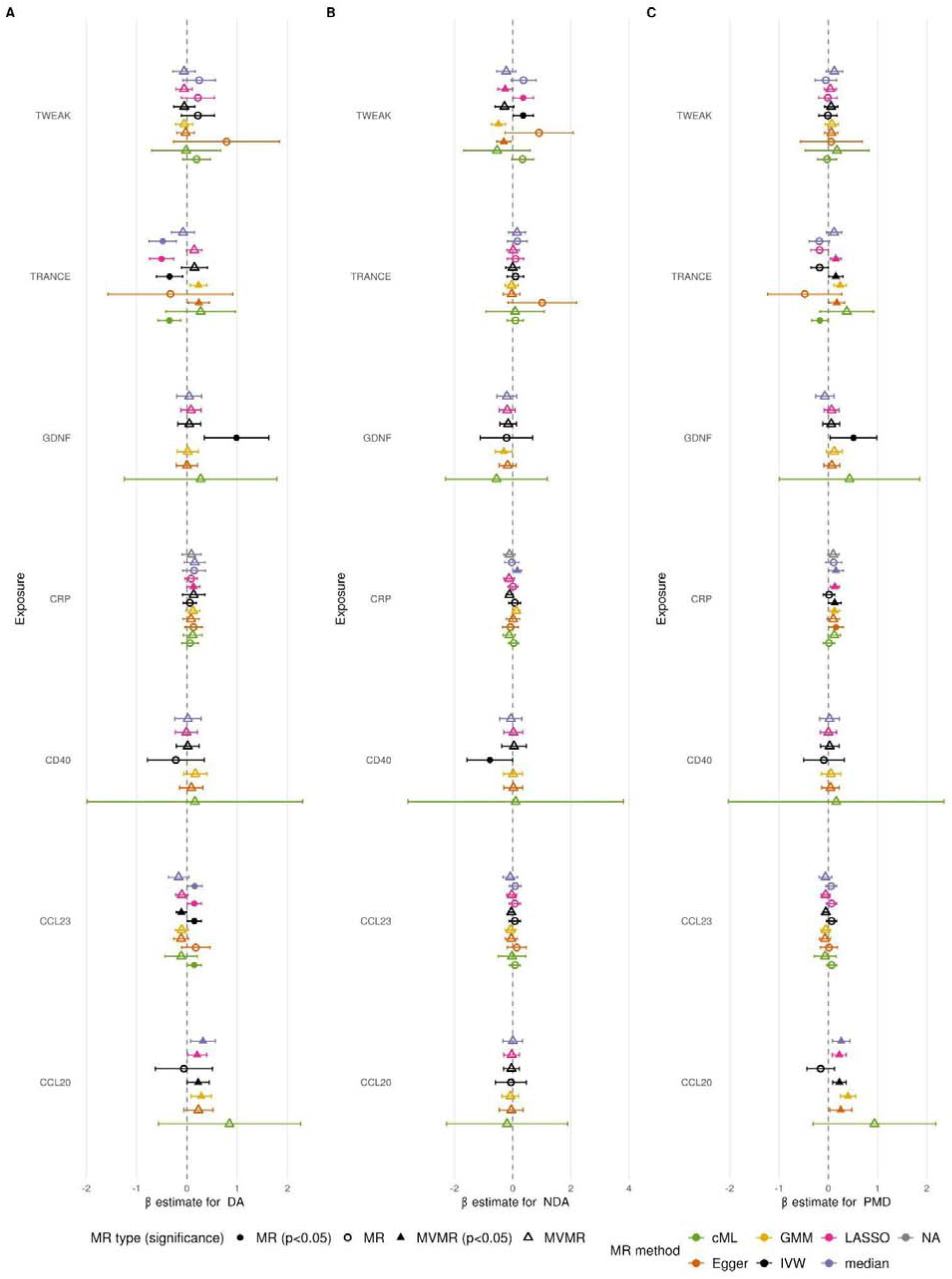
Inverse variance weighted univariable Mendelian randomization (MR) and multivariable MR (MVMR) analysis results for nominally significant associations between circulating inflammatory marker (CIM) instruments with three or more SNPs and mammographic density (MD) phenotypes. CIM instruments with three or more instruments and at least one nominally significant (*p* < 0.05) association with an MD phenotype are listed on the y-axes. MR and MVMR estimates and 95% confidence intervals for A) dense area (DA), B) non-dense area (NDA), and C) percent mammographic density (PMD) are on the x-axes. Estimates for MD outcomes are per 1 SD change in sqrt(%) PMD or sqrt(cm^2^) DA/NDA. Open shapes represent MR or MVMR estimates with *p* ≥ 0.05. Circles represent MR estimates and triangles represent MVMR estimates. Estimates are colored by method (top to bottom: median, LASSO, IVW, GMM, Egger, cML).

**Table 1.**
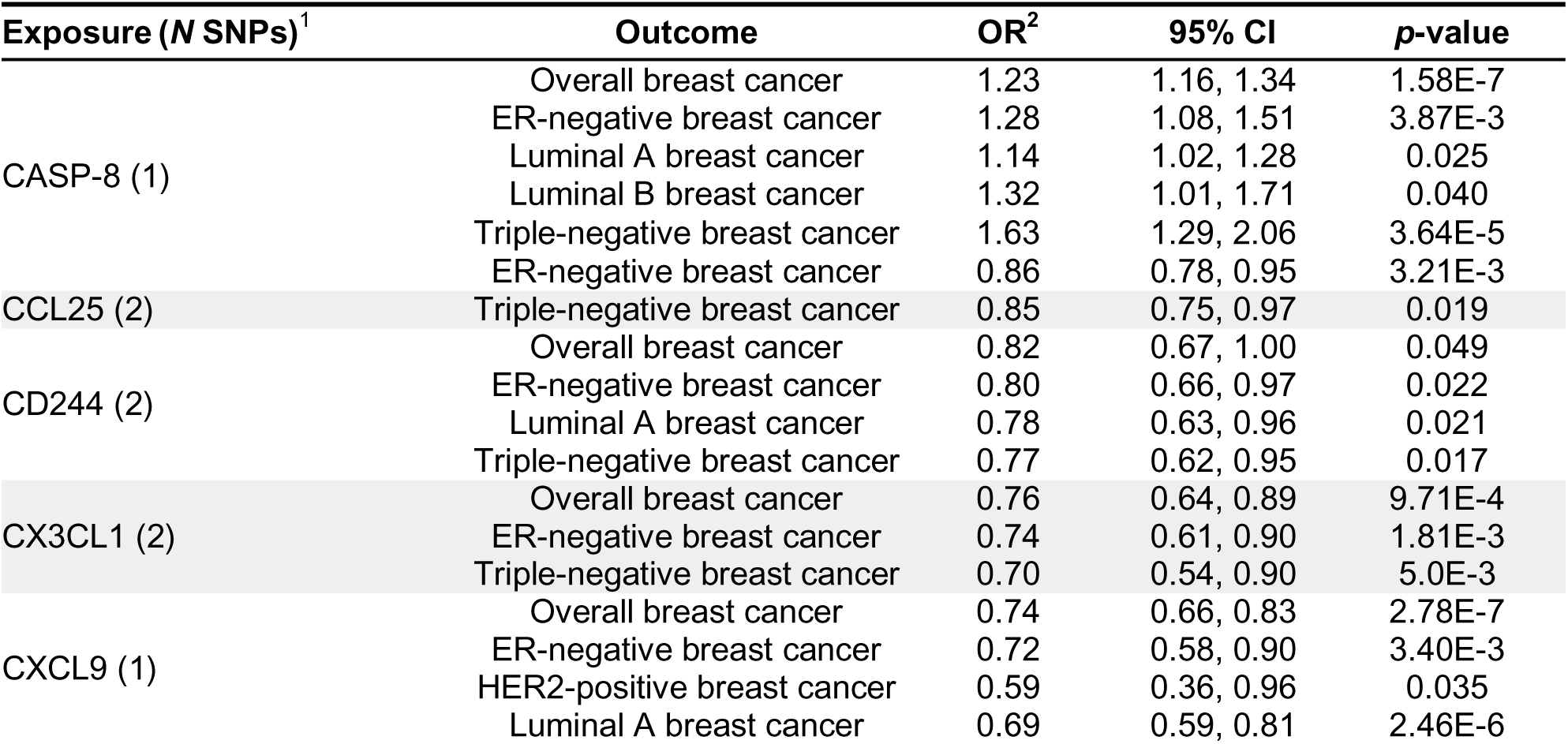
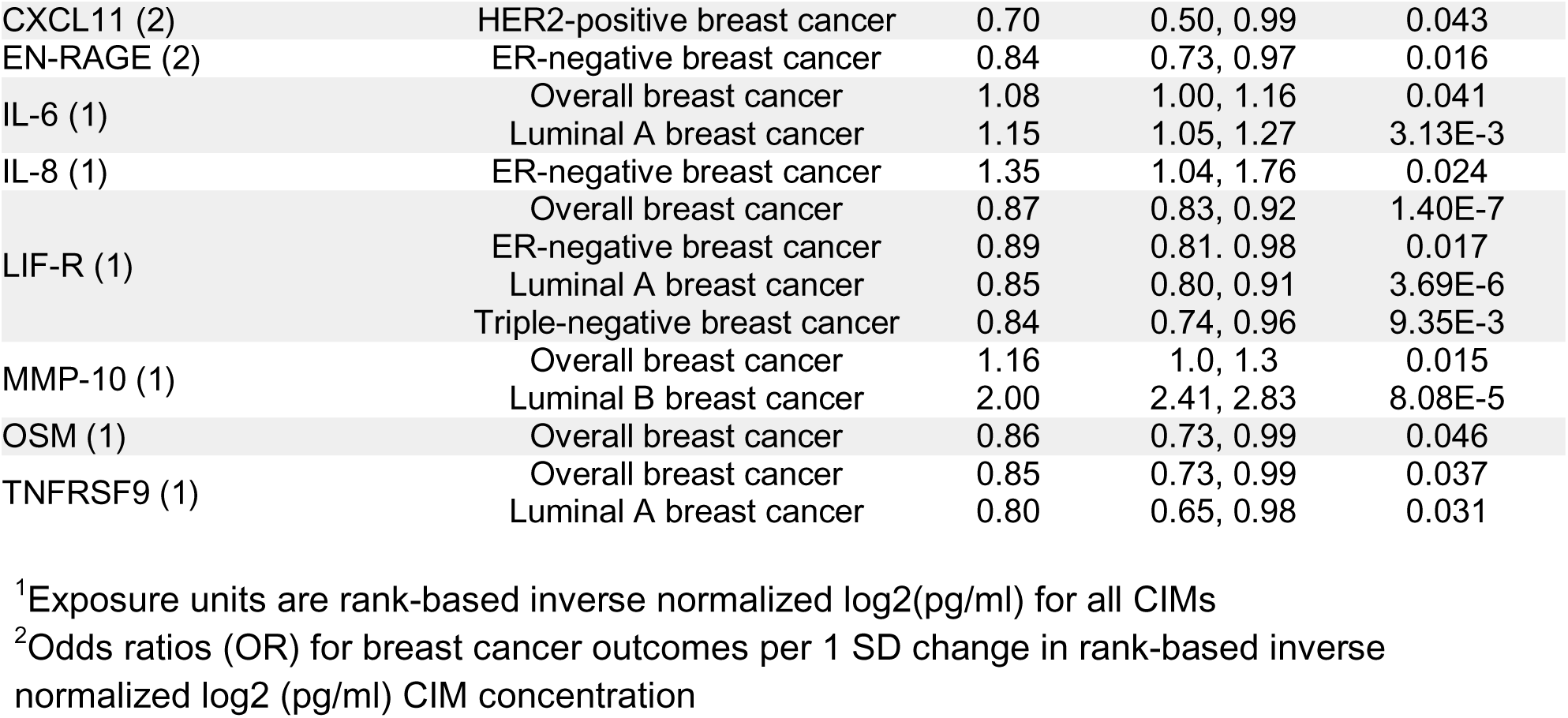
Univariable Mendelian randomization IVW analysis results for breast cancer outcomes with circulating inflammatory markers with two or fewer SNPs that showed nominally significant (*p* < 0.05) associations.

| **Exposure (*N* SNPs)**<sup>1</sup> | **Outcome** | **OR**<sup>2</sup> | **95% CI** | ***p*-value** |
|---|---|---|---|---|
| CASP-8 (1) | Overall breast cancer | 1.23 | 1.16, 1.34 | 1.58E-7 |
|  | ER-negative breast cancer | 1.28 | 1.08, 1.51 | 3.87E-3 |
|  | Luminal A breast cancer | 1.14 | 1.02, 1.28 | 0.025 |
|  | Luminal B breast cancer | 1.32 | 1.01, 1.71 | 0.040 |
|  | Triple-negative breast cancer | 1.63 | 1.29, 2.06 | 3.64E-5 |
|  | ER-negative breast cancer | 0.86 | 0.78, 0.95 | 3.21E-3 |
| CCL25 (2) | Triple-negative breast cancer | 0.85 | 0.75, 0.97 | 0.019 |
| CD244 (2) | Overall breast cancer | 0.82 | 0.67, 1.00 | 0.049 |
|  | ER-negative breast cancer | 0.80 | 0.66, 0.97 | 0.022 |
|  | Luminal A breast cancer | 0.78 | 0.63, 0.96 | 0.021 |
|  | Triple-negative breast cancer | 0.77 | 0.62, 0.95 | 0.017 |
| CX3CL1 (2) | Overall breast cancer | 0.76 | 0.64, 0.89 | 9.71E-4 |
|  | ER-negative breast cancer | 0.74 | 0.61, 0.90 | 1.81E-3 |
|  | Triple-negative breast cancer | 0.70 | 0.54, 0.90 | 5.0E-3 |
| CXCL9 (1) | Overall breast cancer | 0.74 | 0.66, 0.83 | 2.78E-7 |
|  | ER-negative breast cancer | 0.72 | 0.58, 0.90 | 3.40E-3 |
|  | HER2-positive breast cancer | 0.59 | 0.36, 0.96 | 0.035 |
|  | Luminal A breast cancer | 0.69 | 0.59, 0.81 | 2.46E-6 |
| CXCL11 (2) | HER2-positive breast cancer | 0.70 | 0.50, 0.99 | 0.043 |
| EN-RAGE (2) | ER-negative breast cancer | 0.84 | 0.73, 0.97 | 0.016 |
| IL-6 (1) | Overall breast cancer | 1.08 | 1.00, 1.16 | 0.041 |
|  | Luminal A breast cancer | 1.15 | 1.05, 1.27 | 3.13E-3 |
| IL-8 (1) | ER-negative breast cancer | 1.35 | 1.04, 1.76 | 0.024 |
| LIF-R (1) | Overall breast cancer | 0.87 | 0.83, 0.92 | 1.40E-7 |
|  | ER-negative breast cancer | 0.89 | 0.81. 0.98 | 0.017 |
|  | Luminal A breast cancer | 0.85 | 0.80, 0.91 | 3.69E-6 |
|  | Triple-negative breast cancer | 0.84 | 0.74, 0.96 | 9.35E-3 |
| MMP-10 (1) | Overall breast cancer | 1.16 | 1.0, 1.3 | 0.015 |
|  | Luminal B breast cancer | 2.00 | 2.41, 2.83 | 8.08E-5 |
| OSM (1) | Overall breast cancer | 0.86 | 0.73, 0.99 | 0.046 |
| TNFRSF9 (1) | Overall breast cancer | 0.85 | 0.73, 0.99 | 0.037 |
|  | Luminal A breast cancer | 0.80 | 0.65, 0.98 | 0.031 |

<sup>1</sup>Exposure units are rank-based inverse normalized log2(pg/ml) for all CIMs
<sup>2</sup>Odds ratios (OR) for breast cancer outcomes per 1 SD change in rank-based inverse normalized log2 (pg/ml) CIM concentration

Across remaining CIMs, univariable IVW effect estimates for breast cancer and MD phenotype outcomes were generally consistent with those from MR sensitivity analyses for that outcome (Tables S4-S9). Exceptions included were CCL23-ER−/HER2+ breast cancer, CCL28-NDA, CRP-PMD, CD5-overall/HER2+ breast cancer, FGF19-PMD/Luminal B breast cancer, MIP-1-alpha-overall/ER−/HER2+/luminal A breast cancer, TRAIL-ER-breast cancer, and TRANCE-overall/ER−/luminal B breast cancer, which had significant MR-Egger intercepts, indicating possible horizontal pleiotropy (Table S6). The CRP instrument also had significant MR-PRESSO global test p-values in the NDA and PMD analyses, further indicating possible horizontal pleiotropy for those associations (Table S10). We observed limited evidence of horizontal pleiotropy across most univariable MR analyses. No other CIM instruments were nominally associated with any MD outcome in IVW analyses (all *p* ≥ 0.05). MR-LASSO generally flagged few SNPs as potentially invalid across instruments, with only 12 CIMs (CCL11, CCL23, CD5, FGF-19, FGF-21, Flt3L, MCP-4, MIP-1-alpha, SCF, TRAIL, TWEAK, and VEGF-A) having 25% or more of the SNPs in the instrument indicated as potentially invalid for any analysis (Table S8). MR-PRESSO identified eleven potential outlier SNPs (rs6801781, rs113184201, rs704017, rs28429148, rs6090040, rs4821764, rs4817984, rs585406, rs630372, rs73137144, and rs7970695) for the CRP instrument across the analyses (Table S10).

### Multivariable MR analysis

In multivariable MR analyses taking BMI into account, conditional *F*-statistics for all exposures except BMI and CRP were *F* < 10, indicating the potential for weak instrument bias (Table S3). In agreement with our previous work (37), we observed strong inverse associations between BMI and PMD (−0.81 sqrt(%) per 1 kg/m^2^ increase in BMI; 95% CI: −1.02, −0.61; *p* = 1.6 x 10^−15^) and DA (−0.35 sqrt(cm^2^) per 1 kg/m^2^ increase in BMI; 95% CI: −0.60, −0.10; *p* = 0.006), and a positive association between increased BMI and NDA (1.34 sqrt(cm^2^) per 1 kg/m^2^ increase in BMI; 95% CI: 0.95, 1.73; *p* = 1.1 x 10^−11^) in the CRP_BMI multivariable MR analysis. The estimated associations between BMI and MD outcomes were generally consistent across multivariable IVW MR models and similar to those observed in the CRP-BMI multivariable MR analysis (Table S4). We observed nominally significant association between CRP and PMD (0.131 sqrt(%) per 1 ln(mg/l) increase in CRP; 95% CI: 0.00, 0.26; *p* = 0.05), but no evidence of association for DA (*p* = 0.24) or NDA (*p* = 0.06) (Figure 3 & Table S4). We also observed nominally statistically significant associations for CCL23 with DA, TRAIL with NDA, and CCL20, OPG, and TRANCE with PMD, with no evidence of horizontal pleiotropy based on multivariable MR-Egger intercepts (Table S6).

## Discussion

The purpose of this study was to investigate the relationships between genetically predicted CIMs, MD phenotypes, and breast cancer subtypes using an MR approach. In this comprehensive MR analysis of 60 CIMs, we identified 7 significant CIM-breast cancer associations representing 4 unique CIMs. In contrast, we found limited evidence supporting broad causal effects of circulating inflammatory markers on mammographic density phenotypes. Although several nominal associations were observed, none survived correction for multiple testing. Overall, our results suggest that a small subset CIMs may have effects on breast cancer risk but the consistency of largely null findings across the CIMs evaluated suggests that CIMs are unlikely to have broad or strong causal effects on MD phenotypes or breast cancer.

The breast cancer associations we detected in our analyses all represent CIMs with known roles in breast cancer or with associations observed in previous studies. The SNP used for our CASP-8 instrument (rs56328050) is in high LD (r^2^ = 0.99) with a *CASP8* variant (rs1045485) that has known associations with breast cancer risk in prior BCAC studies and other European ancestry population studies (46,53,54). Prior studies have found that CXCL9 expression in breast tumors are correlated with tumor subtype and it has long been considered as a candidate prognostic blood biomarker for breast cancer (55,56). LIF-R and MMP-10 have also been evaluated as a serum biomarkers for breast cancer (57,58). MMP-10 was the only CIM where we identified nominally significant associations with both MD phenotypes (DA and PMD) and breast cancer outcomes (overall and luminal B). This association may warrant further follow up as higher MMP-10 concentrations were associated with higher breast density and increased odds A previous bidirectional MR study of multiple matrix metalloproteinase (MMP) CIMs and ER-negative breast cancer found an association and larger MMP-1 MR instrument that we did not detect despite using similar summary statistics (59). In agreement with our analyses this study also did not observe an association between MMP-10 and ER-negative breast cancer. It should be noted that the MMP-10 variant (rs516316) used in our MR instrument is an intronic *FUT2* variant associated with at least 39 human traits in the GWAS Catalog (36), suggesting a high degree of pleiotropy. While we did not observe direct evidence for pleiotropy from that variant in our analyses, the MMP-10 results should be interpreted with caution. Importantly, we did not observe breast cancer associations for any of the CIMs with previously reported associations for MD phenotypes (IL-6, IL-8, TNFR2, VEGF, or CRP) (21–25).

The nominal associations identified from multi-SNP instruments in univariable MR analyses for MD phenotypes (CCL23, TRANCE, and TWEAK) were highly consistent across all of the MR methods we utilized. With the exception of CRP, none of these CIMs have previously been reported to be associated with MD phenotypes, but all have previously been linked to breast cancer biology, including pathways involving immune activation, apoptosis, and tumor progression (14–17,60–68). However, with the exception of MMP-10, we did not observe any associations with breast cancer subtypes and none of these observed associations were significant after correction for multiple testing. As a result, these findings should be viewed as hypothesis-generating and require replication in independent datasets. Notably, CRP was the only CIM with previously reported observational associations with MD phenotypes for which we observed nominal associations in the present study. We did not identify associations (all *p* ≥ 0.05) between genetically predicted concentrations of IL-6, IL-8, TNFR2, or VEGF and any MD phenotype, suggesting that some previously reported observational associations may reflect confounding, reverse causation, or differences between circulating and local tissue-specific inflammatory processes (22–24).

The association between CCL23 and DA was the only association observed in both the univariable and multivariable MR analyses for MD phenotypes. However, the direction of the observed CCL23-DA association changed from positive in univariable MR to negative in multivariable MR analyses, further reducing confidence that this association reflects a robust causal effect. In addition, the observed negative association for PMD and TRANCE in the multivariable MR analysis conflicts with the observed positive association for DA and TRANCE in the univariable MR analysis (Figures 2 & 3). For all CIM instruments with the exception of CRP, conditional *F* statistics in the multivariable MR analyses indicated weak instruments. In contrast to univariable MR where weak instruments tend to bias estimates towards the null, the direction of bias from weak instruments in multivariable MR is more challenging to predict (41). Thus, these multivariable MR analyses should be interpreted cautiously and considered exploratory.

Our results for CRP were inconsistent across MR methods. In univariable IVW analyses, we observed no associations between CRP and MD phenotypes, whereas in multivariable MR IVW analyses including BMI, we observed a nominal positive association between CRP concentrations and PMD. The magnitude of the CRP-PMD association was generally consistent across the multivariable MR analyses. Overall, we found that inclusion of BMI in multivariable MR analyses reduced conditional instrument strength for most CIMs, highlighting the difficulty of disentangling adiposity-related pathways from systemic inflammatory processes and/or theoretical challenges with multivariable MR when the number of genome-wide significant SNPs varies widely between exposures of interest. Given the strong association between BMI and both CRP and MD phenotypes (4,26–28), it is possible that our multivariable MR results better reflect the underlying relationship between CRP and MD phenotypes, especially as the GWAS summary statistics used to construct CRP instruments were not adjusted for BMI. Indeed, it has been proposed that multivariable MR approaches may produce less biased estimates in the absence of confounder-adjusted exposure or outcome GWAS summary statistics, regardless of the bias correction approaches used for univariable MR (69). Notably, the CRP-PMD association reached nominal significance only in the MVMR analysis, whereas the standard CRP instrument was null and the CRP_noBMI instrument produced a positive but non-significant estimate, suggesting that the estimated direct effect of CRP may be sensitive to how BMI-related genetic variation is accounted for. Further, it is also important to note that previous bidirectional MR studies only estimated the linear effects between BMI and CRP (27,28). Non-linear relationships between BMI and CRP are possible (17), and could make modeling the BMI-CRP relationship in multivariable MR analyses more complex.

While tumor-promoting inflammation is a cancer hallmark (7,8), there is some evidence to suggest that CIMs may not be good proxies for tissue-specific inflammatory markers. An MR study of the relationships between 66 CIMs and 30 adult cancers (including breast cancer) in a European genetic ancestry population found limited evidence that CIM concentrations were associated with cancer (70). While both local and systemic immune responses are relevant to breast cancer risk (71), associations between inflammatory marker concentrations and MD measures have generally been stronger in studies that measured levels in breast tissue (22,23) than in studies that measured CIM levels (21,24,25), suggesting that local inflammation may be more directly relevant to MD. Assessing the relationships between tissue-specific inflammatory markers and MD would therefore be informative given the potential relevance of the breast microenvironment to MD and breast cancer (7,8,22,23). However, we are not aware of studies that provide genetic instruments for breast-specific inflammation markers and were therefore unable to assess these associations here.

Our study has several strengths. To our knowledge, this is the largest and most comprehensive MR analysis of the relationships between CIMs, MD phenotypes, and breast cancer subtypes conducted to date. We applied multiple MR methods with varying degrees of robustness to violations of MR assumptions and used both univariable and multivariable MR approaches to help mitigate the impact of BMI on CRP estimates. We also examined CIM associations with both MD phenotypes and breast cancer subtypes to try and unravel the influences of systemic inflammation in breast cancer risk.

Our study also has some limitations. First, all GWAS were conducted using European genetic ancestry individuals, limiting the generalizability of results to other populations. Given the global importance of breast cancer, examining these relationships in more genetically diverse populations would be informative. In addition, MR results only represent causal effect estimates when all assumptions are satisfied (40,41,49). The presence of pleiotropy or invalid SNPs may violate the assumptions required for causal inference. In the case of our multivariable analyses, we observed the potential for weak instrument bias in the instruments for all CIMs except CRP. While we only observed direct evidence of pleiotropy in our sensitivity analyses for a small number of CIMs, we observed evidence of heterogeneity in a larger number of MR analyses. In addition, several of the identified associations were based on single-SNP instruments, limiting our ability to assess potential horizontal pleiotropy through standard MR sensitivity analyses. We also did not perform an MR mediation analysis to estimate CIM breast cancer effects mediated by MD phenotypes. While this analysis may have provided additional information, we felt that the need to include BMI and multiple MD phenotypes in the mediation MVMR models in order to obtain unbiased estimates would likely diminish our extract useful information. This is because current MVMR mediation analysis approaches do not make it possible to distinguish the contributions of each mediator to the indirect effect (72). Finally, the GWAS for all CIMs except CRP had modest sample sizes (N < 55,000), and larger GWAS may provide more informative IVs.

Overall, this study indicates that a small number of CIMs may be associated with risk of several breast cancer subtypes but provides limited evidence supporting broad causal effects of CIMs on MD phenotypes. These findings suggest that previously reported observational associations between CIMs and MD may not reflect true causal effects, with the possible exception of CRP, for which we observed a small positive association with PMD in multivariable MR analyses. A small number of markers may warrant follow-up, including MMP-10 which showed indications of being associated with both MD phenotypes and breast cancer subtypes. Replication in independent datasets and evaluation using larger GWAS will help determine whether any of the nominally significant signals identified here represent robust causal associations.

## Supporting information

STROBE-MR-checklist

## Acknowledgements

This work was supported by CA244670. A.H.S. was supported by grant number T32CA009168 from the NCI, NIH. The study’s contents are solely the responsibility of the authors and do not necessarily represent the official views of the NCI, NIH. This research was supported [in part] by the Intramural Research Program of the National Institutes of Health (NIH). The contributions of the NIH author(s) are considered Works of the United States Government. The findings and conclusions presented in this paper are those of the author(s) and do not necessarily reflect the views of the NIH or the U.S. Department of Health and Human Services.

## BCAC acknowledgements

We thank all the individuals who took part in these studies and all the researchers, clinicians, technicians, and administrative staff who have enabled this work to be carried out.

ABCFS thanks Maggie Angelakos, Judi Maskiell, and Gillian Dite; BBCS thanks Eileen Williams, Elaine Ryder-Mills, and Kara Sargus.

BCEES thanks Lin Fritschi, Jane Heyworth, and BreastScreen Western Australia.

The BREOGAN study would not have been possible without the contributions of the following: Manuela Gago-Dominguez, Jose Esteban Castelao, Angel Carracedo, Victor Muñoz Garzón, Alejandro Novo Domínguez, Maria Elena Martinez, Sara Miranda Ponte, Carmen Redondo Marey, Maite Peña Fernández, Manuel Enguix Castelo, Maria Torres, Manuel Calaza (BREOGAN), José Antúnez, Máximo Fraga, and the staff of the Department of Pathology and Biobank of the University Hospital Complex of Santiago-CHUS, Instituto de Investigación Sanitaria de Santiago, IDIS, Xerencia de Xestion Integrada de Santiago-SERGAS; Joaquín González-Carreró and the staff of the Department of Pathology and Biobank of University Hospital Complex of Vigo, Instituto de Investigacion Biomedica Galicia Sur, SERGAS, Vigo, Spain.

CBCS thanks study participants, co-investigators, collaborators, staff of the Canadian Breast Cancer Study, and project coordinators Agnes Lai and Celine Morissette.

FHRISK and PROCAS thank NIHR for funding.

kConFab/AOCS wish to thank Heather Thorne, Eveline Niedermayr, all the kConFab research nurses and staff, the heads and staff of the Family Cancer Clinics, and the Clinical Follow-Up Study [which has received funding from the NHMRC, the National Breast Cancer Foundation, Cancer Australia, and the National Institute of Health (USA)] for their contributions to this resource and the many families who contribute to kConFab.

The MCCS was made possible by the contribution of many people, including the original investigators, the teams that recruited the participants and continue working on follow-up and the many thousands of Melbourne residents who continue to participate in the study.

We thank the coordinators, the research staff, and especially the MMHS participants for their continued collaboration on research studies in breast cancer.

The following are NBCS Collaborators: Kristine K. Sahlberg (PhD), Anne-Lise Børresen-Dale (Prof. Em.), Lars Ottestad (MD), Rolf Kåresen (Prof. Em.), Dr. Ellen Schlichting (MD), Marit Muri Holmen (MD), Toril Sauer (MD), Vilde Haakensen (MD), Olav Engebråten (MD), Bjørn Naume (MD), Alexander Fosså (MD), Cecile E. Kiserud (MD), Kristin V. Reinertsen (MD), Åslaug Helland (MD), Margit Riis (MD), Jürgen Geisler (MD), OSBREAC, and Grethe I. Grenaker Alnæs (MSc).

For NHS and NHS2, the study protocol was approved by the Institutional Review Boards of the Brigham and Women’s Hospital and Harvard T.H. Chan School of Public Health and those of participating registries as required. We would like to thank the participants and staff of the NHS and NHS2 for their valuable contributions as well as the following state cancer registries for their help: AL, AZ, AR, CA, CO, CT, DE, FL, GA, ID, IL, IN, IA, KY, LA, ME, MD, MA, MI, NE, NH, NJ, NY, NC, ND, OH, OK, OR, PA, RI, SC, TN, TX, VA, WA, and WY. The authors assume full responsibility for analyses and interpretation of these data.

The OFBCR thanks Gord Glendon, Nayana Weerasooriya, Teresa Selander and the Sinai Health Biospecimen Repository.

PBCS thanks Louise Brinton, Mark Sherman, Neonila Szeszenia-Dabrowska, Beata Peplonska, Witold Zatonski, Pei Chao, and Michael Stagner.

SASBAC thanks the Swedish Medical Research Counsel. We thank the SEARCH and EPIC teams.

UKBGS thanks Breast Cancer Now and the Institute of Cancer Research for support and funding of the Generations Study and the study participants, study staff, and the doctors, nurses, and other health care providers and health information sources who have contributed to the study. We acknowledge NHS funding to the Royal Marsden/ICR NIHR Biomedical Research Center.

## BCAC funding acknowledgements

BCAC is funded by the European Union’s Horizon 2020 Research and Innovation Program (Grant Numbers 634935 and 633784 for BRIDGES and B-CAST, respectively) and the PERSPECTIVE I&I Project, funded by the Government of Canada through Genome Canada and the Canadian Institutes of Health Research, the Ministère de l’Économie et de l’Innovation du Québec through Genome Québec, and the Quebec Breast Cancer Foundation. The EU Horizon 2020 Research and Innovation Program funding source had no role in study design, data collection, data analysis, data interpretation, or writing of the report. Additional funding for BCAC is provided by Cancer Research UK Grant PPRPGM-Nov20/100002 and via the Confluence Project, which is funded with intramural funds from the National Cancer Institute Intramural Research Program, National Institutes of Health.

Genotyping of the OncoArray was funded by the NIH Grant U19 CA148065 and Cancer Research UK Grant C1287/A16563 and the PERSPECTIVE Project supported by the Government of Canada through Genome Canada and the Canadian Institutes of Health Research (Grant GPH-129344), the Ministère de l’Économie, Science et Innovation du Québec through Genome Québec and the PSRSIIRI-701 Grant, and the Quebec Breast Cancer Foundation. Funding for iCOGS came from: the European Community’s Seventh Framework Program under Grant Agreement n° 223175 (HEALTH-F2-2009-223175) (COGS), Cancer Research UK (C1287/A10118, C1287/A10710, C12292/A11174, C1281/A12014, C5047/A8384, C5047/A15007, C5047/A10692, C8197/A16565), the National Institutes of Health (CA128978) and Post-Cancer GWAS Initiative (1U19 CA148537, 1U19 CA148065, and 1U19 CA148112—the GAME-ON Initiative), the Department of Defense (W81XWH-10-1-0341), the Canadian Institutes of Health Research (CIHR) for the CIHR Team in Familial Risks of Breast Cancer, and Komen Foundation for the Cure, the Breast Cancer Research Foundation, and the Ovarian Cancer Research Fund.

The Australian Breast Cancer Family Study (ABCFS) was supported by Grant UM1 CA164920 from the National Cancer Institute (USA). The content of this manuscript does not necessarily reflect the views or policies of the National Cancer Institute or any of the collaborating centers in the Breast Cancer Family Registry (BCFR), nor does mention of trade names, commercial products, or organizations imply endorsement by the USA Government or the BCFR. The ABCFS was also supported by the National Health and Medical Research Council of Australia, the New South Wales Cancer Council, the Victorian Health Promotion Foundation (Australia), and the Victorian Breast Cancer Research Consortium. J.L.H. is a National Health and Medical Research Council (NHMRC) Senior Principal Research Fellow. M.C.S. is a NHMRC Senior Research Fellow.

The work of the BBCC was partly funded by ELAN-Fond of the University Hospital of Erlangen.

The BCEES was funded by the National Health and Medical Research Council, Australia and the Cancer Council Western Australia and acknowledges funding from the National Breast Cancer Foundation (JS).

The BREast Oncology GAlician Network (BREOGAN) is funded by Acción Estratégica de Salud del Instituto de Salud Carlos III FIS PI12/02125/Cofinanciado and FEDER PI17/00918/Cofinanciado FEDER; Acción Estratégica de Salud del Instituto de Salud Carlos III FIS Intrasalud (PI13/01136); Programa Grupos Emergentes, Cancer Genetics Unit, Instituto de Investigacion Biomedica Galicia Sur. Xerencia de Xestion Integrada de Vigo-SERGAS, Instituto de Salud Carlos III, Spain; Grant 10CSA012E, Consellería de Industria Programa Sectorial de Investigación Aplicada, PEME I+D e I+D Suma del Plan Gallego de Investigación, Desarrollo e Innovación Tecnológica de la Consellería de Industria de la Xunta de Galicia, Spain; Grant EC11-192. Fomento de la Investigación Clínica Independiente, Ministerio de Sanidad, Servicios Sociales e Igualdad, Spain; and Grant FEDER-Innterconecta. Ministerio de Economia y Competitividad, Xunta de Galicia, Spain.

CBCS is funded by the Canadian Cancer Society (Grant # 313404) and the Canadian Institutes of Health Research.

FHRISK and PROCAS are funded from NIHR grant PGfAR 0707-10031. DGE, AH and WGN are supported by the NIHR Manchester Biomedical Research Centre (IS-BRC-1215-20007).

kConFab is supported by a Grant from the National Breast Cancer Foundation and previously by the National Health and Medical Research Council (NHMRC), the Queensland Cancer Fund, the Cancer Councils of New South Wales, Victoria, Tasmania and South Australia, and the Cancer Foundation of Western Australia. Financial support for the AOCS was provided by the United States Army Medical Research and Materiel Command (DAMD17-01-1-0729), Cancer Council Victoria, Queensland Cancer Fund, Cancer Council New South Wales, Cancer Council South Australia, The Cancer Foundation of Western Australia, Cancer Council Tasmania, and the National Health and Medical Research Council of Australia (NHMRC; 400413, 400281, 199600). G.C.T. and P.W. are supported by the NHMRC. RB was a Cancer Institute NSW Clinical Research Fellow.

MCBCS was supported by the NIH Grants R35CA253187, R01CA192393, R01CA116167, and R01CA176785, a NIH Specialized Program of Research Excellence (SPORE) in Breast Cancer (P50CA116201), and the Breast Cancer Research Foundation.

The Melbourne Collaborative Cohort Study (MCCS) Cohort Recruitment was funded by VicHealth and Cancer Council Victoria. The MCCS was further augmented by Australian National Health and Medical Research Council Grants 209057, 396414, and 1074383 and by infrastructure provided by Cancer Council Victoria. Cases and their vital status were ascertained through the Victorian Cancer Registry.

The MEC was supported by NIH Grants CA63464, CA54281, CA098758, CA132839, and CA164973.

The MMHS study was supported by NIH Grants CA97396, CA128931, CA116201, CA140286, and CA177150.

The NBCS has received funding from the K.G. Jebsen Center for Breast Cancer Research; the Research Council of Norway Grant 193387/V50 (to A.-L. Børresen-Dale and V.N. Kristensen) and Grant 193387/H10 (to A.-L. Børresen-Dale and V.N. Kristensen), South Eastern Norway Health Authority (Grant 39346 to A.-L. Børresen-Dale), and the Norwegian Cancer Society (to A.-L. Børresen-Dale and V.N. Kristensen).

The Northern California Breast Cancer Family Registry (NC-BCFR) and Ontario Familial Breast Cancer Registry (OFBCR) were supported by Grant U01CA164920 from the USA National Cancer Institute of the National Institutes of Health. The content of this manuscript does not necessarily reflect the views or policies of the National Cancer Institute or any of the collaborating centers in the Breast Cancer Family Registry (BCFR), nor does mention of trade names, commercial products, or organizations imply endorsement by the USA Government or the BCFR.

The NHS was supported by NIH Grants P01 CA87969, UM1 CA186107, R01CA131332, and U19 CA148065. The NHS2 was supported by NIH Grants UM1 CA176726, R01 CA124865, and U19 CA148065.

The PBCS was funded by the Intramural Research Program of the National Cancer Institute, National Institutes of Health, Department of Health and Human Services, USA.

The SASBAC study was supported by funding from the Agency for Science, Technology, and Research of Singapore (A*STAR), the US National Institute of Health (NIH) and the Susan G. Komen Breast Cancer Foundation.

SEARCH is funded by Cancer Research UK (C490/A10124, C490/A16561) and supported by the UK National Institute for Health Research Biomedical Research Center at the University of Cambridge. The University of Cambridge has received salary support for PMDPP from the NHS in the East of England through the Clinical Academic Reserve.

The UKBGS is funded by Breast Cancer Now and the Institute of Cancer Research (ICR), London. ICR acknowledges NHS funding to the NIHR Biomedical Research Center.

## UK Biobank acknowledgement

We are grateful to the UK Biobank participants who have generously agreed to provide a broad range of information for health-related research. This research was conducted using the UK Biobank Resource under Application 95770.

## Data Availability Statement

We used publicly available GWAS summary statistics from individual study, GWAS Catalog, and IEU OpenGWAS project databases for circulating inflammatory markers and BMI as cited. The BCAC data used to generate the mammographic density GWAS summary statistics used in this study are available via application to the Data Access and Co-ordination Committee.

## Code Availability Statement

Code used in these analyses are publicly available in our project GitHub repository (https://github.com/hammea2/MD_CIM_BCa_MR_v2/tree/Final_BCa_analyses).

## Author Contributions

A.H.S.: Conceptualization, Data curation, Formal analysis, Methodology, Visualization, Writing-original draft. T.A.H.: Conceptualization, Data curation, Methodology, Supervision, Writing - original draft. S.Li.: Conceptualization, Funding Acquisition, Methodology, Project administration, Supervision, Writing - original draft. R.M.T and C.M.V.: Funding acquisition, Methodology, Project administration, Resources, Writing - review & editing. C.B.H. and H.C.: Data curation, Formal analysis, Writing - review & editing. G.L.G.: Methodology, Project administration, Resources, Writing - review and editing. M.K.B. and Q.W.: Data curation, Project administration, Resources, Writing - review & editing. S.F., J.D., K.M., A.M.D., D.F.E, C.G.S.: Data curation, Resources, Writing - review & editing. M.C.S., P.A.F., L.H., J.S., M.G-D. J.E.C., J.Ri., R.A.M., DG.E, S.Lo., F.J.C., S.Y., R.L.M., C.A.H, A.N., E.M.J., A.M.M., V.N.K., A.H.E., I.L.A., J.D.F., S.A., K.C. P.D.P.P., A.C.A, M. G-C., and A.B.: Resources, Writing - review & editing.

