## Supplementary material for "Two-sample Mendelian randomization analysis of circulating inflammatory markers, mammographic density phenotypes, and breast cancer subtypes": STROBE-MR-checklist

**STROBE-MR checklist of recommended items to address in reports of Mendelian randomization studies**^1^ ^2^

| **Item No.** | **Section** | **Checklist item** | **Page No.** | **Relevant text from manuscript** |
| --- | --- | --- | --- | --- |
| 1 | **TITLE and ABSTRACT** | Indicate Mendelian randomization (MR) as the study’s design in the title and/or the abstract if that is a main purpose of the study | 1 & 5 | “Title: Circulating inflammatory markers and mammographic density phenotypes: a Mendelian randomization analysis”  “We utilized two-sample Mendelian randomization (MR) to assess relationships between 60 CIMs and MD phenotypes in individuals of European genetic ancestry by examining associations between genetically predicted CIM concentrations and three MD phenotypes (dense area, DA: N = 13,965; non-dense area, NDA: N = 14,036; percent density, PMD: N = 17,841). Primary analyses were conducted using the inverse variance weighted MR method.” |
|  | **INTRODUCTION** |  |  |  |
| 2 | **Background** | Explain the scientific background and rationale for the reported study. What is the exposure? Is a potential causal relationship between exposure and outcome plausible? Justify why MR is a helpful method to address the study question | 6-7 | “Tumor-promoting inflammation is a cancer hallmark [7,8] and circulating inflammatory markers (CIMs) have been associated with breast cancer in molecular and observational epidemiological studies [9–11].”  “Elucidating the relationships between CIMs and breast cancer risk factors could help disentangle the etiological role of systemic inflammation in breast cancer risk. Studies have identified associations between MD phenotypes and circulating concentrations of interleukin 6 (IL-6) [21–23], interleukin 8 (IL-8) [22,23], tumor necrosis factor receptor 2 (TNFR2) [24], vascular endothelial growth factor (VEGF) [23], and CRP [21,22,24,25]. However, similar to the breast cancer studies, many associations were inconsistent across studies and varied by potential confounding factors [21,25].”  “…using a two-sample Mendelian randomization (MR) approach, as MR can help mitigate some methodological limitations of observational studies, such as reverse causation and uncontrolled confounding.” |
| 3 | **Objectives** | State specific objectives clearly, including pre-specified causal hypotheses (if any). State that MR is a method that, under specific assumptions, intends to estimate causal effects | 7 | “The objective of this study was to comprehensively assess associations between genetically predicted CIMs and MD phenotypes using a two-sample Mendelian randomization (MR) approach…” |
|  | **METHODS** |  |  |  |
| 4 | **Study design and data sources** | Present key elements of the study design early in the article. Consider including a table listing sources of data for all phases of the study. For each data source contributing to the analysis, describe the following: |  |  |
|  | a) | Setting: Describe the study design and the underlying population, if possible. Describe the setting, locations, and relevant dates, including periods of recruitment, exposure, follow-up, and data collection, when available. | 8 | “GWAS summary statistics were obtained from either the IEU OpenGWAS [34,35], the EBI-NHGRI GWAS Catalog [36], or study-specific repositories [33], selecting the largest available European genetic ancestry GWAS when multiple were available. For MD phenotypes (DA: N = 13,965; NDA: N = 14,036; PMD: N = 17,841), we used GWAS summary statistics from the Breast Cancer Association Consortium (BCAC) dataset described in Haas et al., 2024 [37] (Table S2).” |
|  | b) | Participants: Give the eligibility criteria, and the sources and methods of selection of participants. Report the sample size, and whether any power or sample size calculations were carried out prior to the main analysis | 8 | “We used summary statistics from published European genetic ancestry GWAS for BMI (N subjects = 461,460) [30], circulating CRP (N = 575,531) [31] and TNFR2 (N = 21,758) [32], and circulating levels of 58 additional inflammation-related proteins (N = 14,824) for which at least one genome-wide significant (p < 5x10-8) single nucleotide polymorphism (SNP) exists [33] (Table S1), for a total of 60 unique CIMs.” |
|  | c) | Describe measurement, quality control and selection of genetic variants | 9-10 | “We used the TwoSampleMR R package v0.6.11 [34], custom functions in R v4.3.2 [40], and PLINK 2.0 alpha v6.5 [41,42] to extract and process exposure GWAS summary statistics, and harmonize the exposure and outcome GWAS summary statistics to create MR instruments. For each exposure GWAS, only SNPs with p < 5x10-8 were extracted, then SNPs with linkage disequilibrium (LD) were excluded (r2 = 0.001 and clumping window = 10,000 base-pairs) using the 1000 Genomes Project European genetic ancestry (1000G EUR) data as an LD reference panel [34,43]. For the multivariable MR instrument, if a SNP was not included in both the CIM and BMI GWAS summary statistics, then a proxy SNP with r2 > 0.8 in the 1000G EUR LD reference panel was used [35]. Ambiguous strand information was resolved using allele frequency information when possible. Palindromic SNPs were retained if strand information could be inferred.  When constructing all instruments, we excluded SNPs with a minor allele frequency (MAF) below 5% in the 1000G EUR reference panel [43].” |
|  | d) | For each exposure, outcome, and other relevant variables, describe methods of assessment and diagnostic criteria for diseases |  | N/A, described in studies that collected these variables |
|  | e) | Provide details of ethics committee approval and participant informed consent, if relevant | 8 | “The original GWAS or studies meta-analyzed by those GWAS obtained ethical approval from their organization/institution(s) ethical review boards as described in the cited articles. The GWAS summary statistics used did not contain any individual-level information.” |
| 5 | **Assumptions** | Explicitly state the three core IV assumptions for the main analysis (relevance, independence and exclusion restriction) as well assumptions for any additional or sensitivity analysis | 9 | “To be valid, instrumental variables (IVs) used in MR analyses must meet three assumptions (MR IV 1-3): (1) the IVs must be strongly associated with the exposure, (2) the IVs must be independent of all (observed or unobserved) confounders of the exposure-outcome relationship, and (3) the IVs must be associated with the outcome only via the exposure (i.e., independent of the outcome given the exposure; Figure 1A) [38]. The expanded assumptions (multivariable MR IV 1-3) for valid IVs in multivariable MR analyses are that: (1) the IVs must be strongly associated with each exposure given all other exposures in the model, (2) the IVs must be independent of all confounders of any of the relationships between the exposures and the outcome, and (3) the IVs must be independent of the outcome given all exposures (Figure 1B) [38,39].” |
| 6 | **Statistical methods: main analysis** | Describe statistical methods and statistics used |  |  |
|  | a) | Describe how quantitative variables were handled in the analyses (i.e., scale, units, model) | 24 + Tables | “^1^Exposure units were ln(mg/l) for CRP and rank-based inverse normalized log2 (pg/ml) for all other CIMs  ^2^Outcome units were sqrt(cm2) for DA and NDA and sqrt(%) for PMD”  Supplementary table 1 include information on the units and GWAS models for all instruments used. |
|  | b) | Describe how genetic variants were handled in the analyses and, if applicable, how their weights were selected | 10 | “For all CIM instruments, we conducted univariable MR and multivariable MR analyses using random-effect inverse-variance weighted (IVW) methods [45]. These methods were implemented in the MendelianRandomization R package v0.10.0 [46].” |
|  | c) | Describe the MR estimator (e.g. two-stage least squares, Wald ratio) and related statistics. Detail the included covariates and, in case of two-sample MR, whether the same covariate set was used for adjustment in the two samples | 10 + Tables | “Primary inference was based on IVW MR analyses, with nominal p-values providing suggestive evidence in the context of multiple testing across CIM-MD phenotype pairs.”  Supplementary table 1 include information on the GWAS models for all instruments used. |
|  | d) | Explain how missing data were addressed | 11 | N/A, only summary statistics were used in our analyses. The original manuscripts that generated the summary statistics describe the handling of missing data where applicable. |
|  | e) | If applicable, indicate how multiple testing was addressed |  | “A total of 61 CIM instruments with at least one valid SNP were tested using the univariable MR method for each of the three MD outcomes. CRP had two instruments for univariable MR analyses as described above. This resulted in 183 (61 x 3) total univariable MR analyses representing 180 (60 x 3) unique CIM-MD phenotype relationships. For multivariable MR, a total of 60 instruments resulted in 180 total multivariable MR analyses for 180 unique CIM-MD associations. As a result, the corresponding Bonferroni-corrected significance thresholds were p < 2.73 × 10-4 (0.05 / 183) and p < 2.78 × 10-4 (0.05 / 180), respectively, for univariable and multivariable IVW MR analyses.” |
| 7 | **Assessment of assumptions** | Describe any methods or prior knowledge used to assess the assumptions or justify their validity | 7 | “We further performed multivariable MR analyses including genetically-predicted BMI as an exposure, given its strong association with MD phenotypes and involvement in inflammatory processes [4,26]. Previous bidirectional MR studies of CRP and BMI have suggested that BMI is likely to affect CRP, rather than vice versa, and thus BMI is a potential confounder in CRP-MD phenotype relationships [27,28]” |
| 8 | **Sensitivity analyses and additional analyses** | Describe any sensitivity analyses or additional analyses performed (e.g. comparison of effect estimates from different approaches, independent replication, bias analytic techniques, validation of instruments, simulations) | 11-12 | “In univariable MR analyses, we calculated F-statistics for each of the instruments to assess the MR IV1 assumption [46]. Although there are no formal tests for the MR IV2 and IV3 assumptions, we used up to seven methods with varying degrees of robustness to help identify potential violations. For instruments with at least one SNP, we calculated MR Wald ratios for each SNP in the instrument. For instruments with more than two SNPs, we further used the MR-Egger [47], MR weighted median (MR median), MR-LASSO [48], and MR constrained maximum-likelihood (MR cML) [49] methods implemented in the MendelianRandomization R package [46]. For instruments with more than 10 SNPs, we used the MR pleiotropy residual sum and outlier (MR-PRESSO) method [50]. The MR-LASSO, MR cML, and MR-PRESSO methods support the identification of outliers and potentially invalid SNPs in the instruments.  To assess the multivariable MR IV1 assumption, we calculated Sanderson‐Windmeijer conditional F-statistics for each exposure in the multivariable MR analyses. For the CRP multivariable MR analyses, we also incorporated the estimated phenotypic correlation between CRP and BMI in UK Biobank (UKB) participants into the conditional F-statistic calculations. This phenotypic correlation was estimated by Pearson correlation between CRP ln(mg/l) and BMI (kg/m2) at baseline for 466,356 participants with available data. To identify violations of multivariable MR IV2-IV3 and outliers or invalid SNPs, we used multivariable extensions of the MR-Egger, MR weighted median, MR-LASSO, MR-cML, and MR generalized method of moments (MR GMM) methods implemented in the MendelianRandomization R package [34]. For MR cML and multivariable MR cML, the range of potentially invalid IVs was based on the number of potentially invalid IVs detected by MR-LASSO or multivariable MR-LASSO, respectively. Given the strong correlation between BMI and CRP, we used two additional multivariable MR methods for the CRP and BMI (CRP_BMI) analyses: the multivariable MR-PRESSO method implemented in the MR-PRESSO R package [50] and robust Q-statistic minimization method implemented in the multivariable MR (MVMR) R package v0.4.1 [39].” |
| 9 | **Software and pre-registration** |  |  |  |
|  | a) | Name statistical software and package(s), including version and settings used | 9-12 | “TwoSampleMR R package v0.6.11 [34]  custom functions in R v4.3.2 [40]  PLINK 2.0 alpha v6.5 [41,42]  MendelianRandomization R package v0.10.0 [46]  MR pleiotropy residual sum and outlier (MR-PRESSO) method [50]  robust Q-statistic minimization method implemented in the multivariable MR (MVMR) R package v0.4.1 [39]” |
|  | b) | State whether the study protocol and details were pre-registered (as well as when and where) | N/A |  |
|  | **RESULTS** |  |  |  |
| 10 | **Descriptive data** |  |  |  |
|  | a) | Report the numbers of individuals at each stage of included studies and reasons for exclusion. Consider use of a flow diagram | N/A | No exclusions of specific individuals were made. As described in the methods, we selected summary statistics sets based on GWAS outcome, sample size, and availability of data. We excluded summary statistics for particular markers when no SNPs showed genome-wide significant (p < 5E-8) associations with the marker. |
|  | b) | Report summary statistics for phenotypic exposure(s), outcome(s), and other relevant variables (e.g. means, SDs, proportions) | Tables | Summary statistics are available for MD outcomes and reported in the supplementary tables. Summary information about inflammatory marker exposures are available in the cited manuscripts that generated the data. |
|  | c) | If the data sources include meta-analyses of previous studies, provide the assessments of heterogeneity across these studies | N/A | These are provided in the referenced studies that generated the data. |
|  | d) | For two-sample MR:  i.  Provide justification of the similarity of the genetic variant-exposure associations between the exposure and outcome samples  ii.  Provide information on the number of individuals who overlap between the exposure and outcome studies | 12 | “The SNP-exposure effects estimated in the CIM GWAS populations are expected to be similar to those in the MD GWAS population, as all individuals were of inferred European genetic ancestry. None of the BCAC studies included in the MD GWASs were included in any of the CIM GWASs, and any sample overlap is deemed minimal (Tables S1 & S2).” |
| 11 | **Main results** |  |  |  |
|  | a) | Report the associations between genetic variant and exposure, and between genetic variant and outcome, preferably on an interpretable scale | 13-14 | “All the SNPs included in the CIM instruments were either associated with that CIM or were in high LD (r2 > 0.8) with a SNP associated with that CIM in the GWAS Catalog or a previous publication [36,44].  In agreement with our previous work [37], we observed strong inverse associations between BMI and PMD (-0.81 sqrt(%) per 1 kg/m2 increase in BMI; 95% CI: -1.02, -0.61; p = 1.6 x 10-15) and DA (-0.35 sqrt(cm2) per 1 kg/m2 increase in BMI; 95% CI: -0.60, -0.10; p = 0.006), and a positive association between increased BMI and NDA (1.34 sqrt(cm2) per 1 kg/m2 increase in BMI; 95% CI: 0.95, 1.73; p = 1.1 x 10-11) in the CRP_BMI multivariable MR analysis. The estimated associations between BMI and MD outcomes were generally consistent across multivariable IVW MR models and similar to those observed in the CRP-BMI multivariable MR analysis (Table S4). We observed nominally significant association between CRP and PMD (0.131 sqrt(%) per 1 ln(mg/l) increase in CRP; 95% CI: 0.00, 0.26; p = 0.05), but no evidence of association for DA (p = 0.24) or NDA (p = 0.06) (Figure 3 & Table S4).”  More details are included in figures and supplementary tables. |
|  | b) | Report MR estimates of the relationship between exposure and outcome, and the measures of uncertainty from the MR analysis, on an interpretable scale, such as odds ratio or relative risk per SD difference | 14 | “In agreement with our previous work [37], we observed strong inverse associations between BMI and PMD (-0.81 sqrt(%) per 1 kg/m2 increase in BMI; 95% CI: -1.02, -0.61; p = 1.6 x 10-15) and DA (-0.35 sqrt(cm2) per 1 kg/m2 increase in BMI; 95% CI: -0.60, -0.10; p = 0.006), and a positive association between increased BMI and NDA (1.34 sqrt(cm2) per 1 kg/m2 increase in BMI; 95% CI: 0.95, 1.73; p = 1.1 x 10-11) in the CRP_BMI multivariable MR analysis. The estimated associations between BMI and MD outcomes were generally consistent across multivariable IVW MR models and similar to those observed in the CRP-BMI multivariable MR analysis (Table S4). We observed nominally significant association between CRP and PMD (0.131 sqrt(%) per 1 ln(mg/l) increase in CRP; 95% CI: 0.00, 0.26; p = 0.05), but no evidence of association for DA (p = 0.24) or NDA (p = 0.06) (Figure 3 & Table S4).”  More details are included in figures and supplementary tables. |
|  | c) | If relevant, consider translating estimates of relative risk into absolute risk for a meaningful time period | N/A | All exposures and outcomes were measurements of biomarker concentrations, adiposity (BMI), or tissue composition (MD outcomes). |
|  | d) | Consider plots to visualize results (e.g. forest plot, scatterplot of associations between genetic variants and outcome versus between genetic variants and exposure) | 14 & 16 | Forest plots for MR IVW primary analyses and sensitivity analysis MR methods are included as main text figures. |
| 12 | **Assessment of assumptions** |  |  |  |
|  | a) | Report the assessment of the validity of the assumptions | 13-16 | “All instruments used in univariable MR analyses had F-statistics > 10, suggesting strong instruments (Table S3)  Across remaining CIMs, univariable IVW effect estimates were generally consistent with those from MR sensitivity analyses (Tables S4-S9). Exceptions were CCL28-NDA, CRP-PMD, and FGF19-PMD, which had significant MR-Egger intercepts, indicating possible horizontal pleiotropy (Table S6). The CRP instrument also had significant MR-PRESSO global test p-values in the NDA and PMD analyses, further indicating possible horizontal pleiotropy for those associations (Table S10). We observed limited evidence of horizontal pleiotropy across most univariable MR analyses. No other CIM instruments were nominally associated with any MD outcome in IVW analyses (all p ≥ 0.05). MR-LASSO generally flagged few (0 to 24) SNPs as potentially invalid across instruments (Table S8). MR-PRESSO identified three potential outlier SNPs (rs28429148 and rs4817984) for the CRP instrument in the NDA analysis, two potential outlier SNPs (rs2393794 and rs4817984) for the CRP_noBMI instrument in the NDA analysis, and one outlier SNP (rs28429148) for the CRP instrument in the PMD analysis (Table S10).  The association between CCL23 and DA was the only association observed in both the univariable and multivariable MR analyses. However, the direction of the observed CCL23-DA association changed from positive in univariable MR to negative in multivariable MR analyses, further reducing confidence that this association reflects a robust causal effect. In addition, the observed negative association for PMD and TRANCE in the multivariable MR analysis conflicts with the observed positive association for DA and TRANCE in the univariable MR analysis (Figures 2 & 3). For all CIM instruments with the exception of CRP, conditional *F* statistics in the multivariable MR analyses indicated weak instruments. In contrast to univariable MR where weak instruments tend to bias estimates towards the null, the direction of bias from weak instruments in multivariable MR is more challenging to predict (39). Thus, these multivariable MR analyses should be interpreted cautiously and considered exploratory.” |
|  | b) | Report any additional statistics (e.g., assessments of heterogeneity across genetic variants, such as *I^2^*, Q statistic or E-value) | Tables | Additional information is provided in the supplementary tables. |
| 13 | **Sensitivity analyses and additional analyses** |  |  |  |
|  | a) | Report any sensitivity analyses to assess the robustness of the main results to violations of the assumptions | 13-16 | “All instruments used in univariable MR analyses had F-statistics > 10, suggesting strong instruments (Table S3)  Across remaining CIMs, univariable IVW effect estimates were generally consistent with those from MR sensitivity analyses (Tables S4-S9). Exceptions were CCL28-NDA, CRP-PMD, and FGF19-PMD, which had significant MR-Egger intercepts, indicating possible horizontal pleiotropy (Table S6). The CRP instrument also had significant MR-PRESSO global test p-values in the NDA and PMD analyses, further indicating possible horizontal pleiotropy for those associations (Table S10). We observed limited evidence of horizontal pleiotropy across most univariable MR analyses. No other CIM instruments were nominally associated with any MD outcome in IVW analyses (all p ≥ 0.05). MR-LASSO generally flagged few (0 to 24) SNPs as potentially invalid across instruments (Table S8). MR-PRESSO identified three potential outlier SNPs (rs28429148 and rs4817984) for the CRP instrument in the NDA analysis, two potential outlier SNPs (rs2393794 and rs4817984) for the CRP_noBMI instrument in the NDA analysis, and one outlier SNP (rs28429148) for the CRP instrument in the PMD analysis (Table S10).  The association between CCL23 and DA was the only association observed in both the univariable and multivariable MR analyses. However, the direction of the observed CCL23-DA association changed from positive in univariable MR to negative in multivariable MR analyses, further reducing confidence that this association reflects a robust causal effect. In addition, the observed negative association for PMD and TRANCE in the multivariable MR analysis conflicts with the observed positive association for DA and TRANCE in the univariable MR analysis (Figures 2 & 3). For all CIM instruments with the exception of CRP, conditional *F* statistics in the multivariable MR analyses indicated weak instruments. In contrast to univariable MR where weak instruments tend to bias estimates towards the null, the direction of bias from weak instruments in multivariable MR is more challenging to predict (39). Thus, these multivariable MR analyses should be interpreted cautiously and considered exploratory.” |
|  | b) | Report results from other sensitivity analyses or additional analyses | Tables | Complete results from all MR and MVMR sensitivity analyses are included in the supplementary tables. |
|  | c) | Report any assessment of direction of causal relationship (e.g., bidirectional MR) | N/A |  |
|  | d) | When relevant, report and compare with estimates from non-MR analyses | N/A |  |
|  | e) | Consider additional plots to visualize results (e.g., leave-one-out analyses) | N/A |  |
|  | **DISCUSSION** |  |  |  |
| 14 | **Key results** | Summarize key results with reference to study objectives | 15 | The purpose of this study was to investigate the relationships between genetically predicted CIMs and MD phenotypes using an MR approach. In this comprehensive MR analysis of 60 CIMs, we found limited evidence supporting broad causal effects of circulating inflammatory markers on mammographic density phenotypes. Although several nominal associations were observed, none survived correction for multiple testing. Overall, the consistency of largely null findings across the CIMs evaluated suggests that circulating inflammatory markers are unlikely to have broad or strong causal effects on MD phenotypes. The nominal associations identified from multi-SNP instruments in univariable MR analyses (CCL23, TRANCE, and TWEAK) were highly consistent across all of the MR methods we utilized. With the exception of CRP, none of these CIMs have previously been reported to be associated with MD phenotypes, but all have previously been linked to breast cancer biology, including pathways involving immune activation, apoptosis, and tumor progression [[14–17,51–59]](https://www.zotero.org/google-docs/?FAV8wc).  We did not identify associations (all p > 0.05) between genetically predicted concentrations of IL-6, IL-8, TNFR2, or VEGF and any MD phenotype, suggesting that some previously reported observational associations may reflect confounding, reverse causation, or differences between circulating and local tissue-specific inflammatory processes [22–24].” |
| 15 | **Limitations** | Discuss limitations of the study, taking into account the validity of the IV assumptions, other sources of potential bias, and imprecision. Discuss both direction and magnitude of any potential bias and any efforts to address them | 15-19 | “However, because none of these observed associations were significant after multiple testing correction, these findings should be viewed as hypothesis-generating and require replication in independent datasets.  The association between CCL23 and DA was the only association observed in both the univariable and multivariable MR analyses. However, the direction of the observed CCL23-DA association changed from positive in univariable MR to negative in multivariable MR analyses, further reducing confidence that this association reflects a robust causal effect. In addition, the observed negative association for PMD and TRANCE in the multivariable MR analysis conflicts with the observed positive association for DA and TRANCE in the univariable MR analysis (Figures 2 & 3). For all CIM instruments with the exception of CRP, conditional F statistics in the multivariable MR analyses indicated weak instruments. In contrast to univariable MR where weak instruments tend to bias estimates towards the null, the direction of bias from weak instruments in multivariable MR is more challenging to predict (39). Thus, these multivariable MR analyses should be interpreted cautiously and considered exploratory.  Our study also has some limitations. First, all GWAS were conducted using European genetic ancestry individuals, limiting the generalizability of results to other populations. Given the global importance of breast cancer, examining these relationships in more genetically diverse populations would be informative. In addition, MR results only represent causal effect estimates when all assumptions are satisfied [[38,39,47]](https://www.zotero.org/google-docs/?MryE7w). The presence of pleiotropy or invalid SNPs may violate the assumptions required for causal inference. In the case of our multivariable analyses, we observed the potential for weak instrument bias in the instruments for all CIMs except CRP. While we only observed direct evidence of pleiotropy in our sensitivity analyses for a small number of CIMs, we observed evidence of heterogeneity in a larger number of MR analyses. In addition, several of the identified associations were based on single-SNP instruments, limiting our ability to assess potential horizontal pleiotropy through standard MR sensitivity analyses. Finally, the GWAS for all CIMs except CRP had modest sample sizes (N < 55,000), and larger GWAS may provide more informative IVs.” |
| 16 | **Interpretation** |  |  |  |
|  | a) | Meaning: Give a cautious overall interpretation of results in the context of their limitations and in comparison with other studies | 19 | “Overall, this study provides limited evidence supporting broad causal effects of circulating inflammatory markers on MD phenotypes. These findings suggest that previously reported observational associations between CIMs and MD may not reflect true causal effects, with the possible exception of CRP, for which we observed a small positive association with PMD in multivariable MR analyses. A small number of markers may warrant follow-up. Replication in independent datasets and evaluation using larger GWAS will help determine whether any of the nominally significant signals identified here represent robust causal associations.” |
|  | b) | Mechanism: Discuss underlying biological mechanisms that could drive a potential causal relationship between the investigated exposure and the outcome, and whether the gene-environment equivalence assumption is reasonable. Use causal language carefully, clarifying that IV estimates may provide causal effects only under certain assumptions | 15-18 | “The nominal associations identified from multi-SNP instruments in univariable MR analyses (CCL23, TRANCE, and TWEAK) were highly consistent across all of the MR methods we utilized. With the exception of CRP, none of these CIMs have previously been reported to be associated with MD phenotypes, but all have previously been linked to breast cancer biology, including pathways involving immune activation, apoptosis, and tumor progression [14–17,51–59]  Further, it is also important to note that previous bidirectional MR studies only estimated the linear effects between BMI and CRP [27,28]. Non-linear relationships between BMI and CRP are possible [17], and could make modeling the BMI-CRP relationship in multivariable MR analyses more complex.  An MR study of the relationships between 66 CIMs and 30 adult cancers (including breast cancer) in a European genetic ancestry population found limited evidence that CIM concentrations were associated with cancer [61]. While both local and systemic immune responses are relevant to breast cancer risk [62], associations between inflammatory marker concentrations and MD measures have generally been stronger in studies that measured levels in breast tissue [22,23] than in studies that measured CIM levels [21,24,25], suggesting that local inflammation may be more directly relevant to MD. Assessing the relationships between tissue-specific inflammatory markers and MD would therefore be informative given the potential relevance of the breast microenvironment to MD and breast cancer [7,8,22,23]. However, we are not aware of studies that provide genetic instruments for breast-specific inflammation markers and were therefore unable to assess these associations here.” |
|  | c) | Clinical relevance: Discuss whether the results have clinical or public policy relevance, and to what extent they inform effect sizes of possible interventions | NA | Given that the results generally do not support broad causal effects of circulating inflammatory markers on MD outcomes, there are no immediate clinical or public policy implications. |
| 17 | **Generalizability** | Discuss the generalizability of the study results (a) to other populations, (b) across other exposure periods/timings, and (c) across other levels of exposure | 18 | “First, all GWAS were conducted using European genetic ancestry individuals, limiting the generalizability of results to other populations. Given the global importance of breast cancer, examining these relationships in more genetically diverse populations would be informative.” |
|  | **OTHER INFORMATION** |  |  |  |
| 18 | **Funding** | Describe sources of funding and the role of funders in the present study and, if applicable, sources of funding for the databases and original study or studies on which the present study is based | 26-31 | Sources of funding for the present study and BCAC are included in the acknowledgements. |
| 19 | **Data and data sharing** | Provide the data used to perform all analyses or report where and how the data can be accessed, and reference these sources in the article. Provide the statistical code needed to reproduce the results in the article, or report whether the code is publicly accessible and if so, where | 31-32 | We used publicly available GWAS summary statistics from individual study, GWAS Catalog, and IEU OpenGWAS project databases for circulating inflammatory markers and BMI as cited. The BCAC data used to generate the mammographic density GWAS summary statistics used in this study are available via application to the Data Access and Co-ordination Committee.  Code used in these analyses are publicly available in our project GitHub repository (<https://github.com/UW-Epidemiology/MD_CIM_MR>) |
| 20 | **Conflicts of Interest** | All authors should declare all potential conflicts of interest | 3-4 | P.A.F. conducts research funded by Amgen, Novartis and Pfizer and received Honoraria from Roche, Novartis and Pfizer. R.A.M. is a consultant for Pharmavite. S.Lo. receives research funding to institution from Novartis, Bristol Myers Squibb, Astra Zeneca/Daiichi Sankyo, Roche-Genentech, MSD, Pfizer, Gilead Sciences, Nektar Therapeutics and Eli Lilly and has acted as consultant to Roche-Genentech, MSD, Gilead Sciences, Astra Zeneca/Daiichi Sankyo, Bristol Myers Squibb, Novartis, Eli Lilly, Amaroq Therapeutics, Mersana Therapeutics, Domain Therapeutics, BioNTech, Bicycle Therapeutics, Exact Sciences, Menari Asia-Pacific, SAGA Diagnostics, Adanate. |

This checklist is copyrighted by the Equator Network under the Creative Commons Attribution 3.0 Unported (CC BY 3.0) license.

1.	Skrivankova VW, Richmond RC, Woolf BAR, Yarmolinsky J, Davies NM, Swanson SA, et al. Strengthening the Reporting of Observational Studies in Epidemiology using Mendelian Randomization (STROBE-MR) Statement. JAMA. 2021;under review.

2.	Skrivankova VW, Richmond RC, Woolf BAR, Davies NM, Swanson SA, VanderWeele TJ, et al. Strengthening the Reporting of Observational Studies in Epidemiology using Mendelian Randomisation (STROBE-MR): Explanation and Elaboration. BMJ. 2021;375:n2233.
